# Critical COVID-19 represents an endothelial disease with high similarity to kidney disease on the molecular level

**DOI:** 10.1101/2021.02.22.21252207

**Authors:** Justyna Siwy, Ralph Wendt, Amaya Albalat, Tianlin He, Harald Mischak, William Mullen, Agnieszka Latosińska, Christoph Lübbert, Sven Kalbitz, Alexandre Mebazaa, Björn Peters, Bernd Stegmayr, Goce Spasovski, Thorsten Wiech, Jan A. Staessen, Johannes Wolf, Joachim Beige

**Author notes:** Corresponding author: Dr. Justyna Siwy, Mosaiques diagnostics GmbH, Rotenburger Straße 20, D-30659 Hannover, GERMANY. These authors contributed equally to this work (Justyna Siwy, Ralph Wendt).

## Abstract

In patients with critical or mild COVID19 (WHO stages 6-8 [n=53] and stages 1-3 [n=66]), 593 urinary peptides significantly affected by disease severity were identified, reflecting the molecular pathophysiology associated with the course of the infection. The peptide profiles were similar compared with those observed in kidney disease, a prototype of target organ damage with major microvascular involvement, thereby confirming the observation that endothelial damage is a hallmark of COVID19. The clinical corollary is that COVID19 is an indication for anti-oxidative, anti-inflammatory and immunosuppressive treatment modalities protecting the endothelial lining.

## Introduction

The pathogenesis of coronavirus disease (COVID-19) caused by the severe acute respiratory syndrome coronavirus 2 (SARS-CoV-2) is still not completely understood. Accumulating evidence indicates that endothelial injury and subsequent capillary leakage with cytokine release and pathological sequelae may be a key factor. In several recent studies renal involvement in COVID-19 was reported [1-3] with special consideration of chronic kidney disease (CKD) as key risk factor for hospitalisation and mortality[4]. CKD is known to be related to endothelial dysfunction [5].

Based on these and additional findings, we hypothesized that severe COVID-19 may be a microvascular disease, displaying similarity to CKD and other diseases such as induced by hanta virus [6], thrombotic microangiopathy and severe sepsis with multi organ dysfunction [7]. Most of these diseases are associated with acute kidney injury (AKI) that may progress to CKD [7]. Pathophysiological mechanisms of COVID-19, especially those linked to protein degradation are expected to be detectable, possibly even enriched in urine peptides.

To test this hypothesis, we performed a comprehensive analysis of urinary peptides of COVID-19 patients using capillary electrophoresis-coupled mass spectrometry (CE-MS) and compared the data from patients with critical and mild courses of COVID-19. In a second step, we used additional proteomics data retrieved from our database [8] and compared the findings to the changes observed in non-COVID-19 patients with diabetic kidney disease (DKD) as a leading cause of CKD, AKI, and heart failure (HF) versus controls.

## Methods

### Patients

Urine samples from 119 SARS-CoV-2 infected patients were collected within a prospective study named Prospective Validation of a Proteomic Urine Test for Early and Accurate Prognosis of Critical Course Complications in Patients with SARS-CoV-2 Infection (CRIT-Cov-U), described previously [9]. Samples were collected within 2 days after a positive real-time reverse transcriptase-polymerase chain reaction test. Disease severity was classified based on the World Health Organisation (WHO)[10] scale as mild (WHO stages 1-3), intermediate (WHO stages 4-5), or critical (WHO stages 6-8). No significant differences in kidney function between the mild and critical group were observed. This study was approved by the local ethics committee (Saxon Chamber of Physicians, registry number #EK-BR-88/20-1), with a waiver of informed consent and by the Institutional Review Boards of all participating centres.

### Proteomics

Samples were analysed by CE-MS and the generated data processed as described before, including assignment of sequences [8;11]

### Flow cytometry

Fluorescence-activated cell sorting (FACS) was performed for COVID-19 patients with normal lymphocyte count (WHO stage 1-4, n=6), COVID-19 patients with lymphopenia (WHO stage 5-8, n=12), and healthy staff members (n=10). Further, we included human immunodeficiency virus (HIV) infected patients with (n=17) and without lymphopenia (n=5) as control to exclude non-COVID-19 lymphopenia effects.

Initially, lymphocyte counts were measured on a particle counter (Sysmex XN-2000, Norderstedt, Germany). The percentage values of studied subsets obtained by cytometric reading were converted into absolute values according to absolute number of lymphocytes count. Immunophenotyping of lymphocytes was performed by an 8-color T (CD3+), and B (CD19+) cytometry with monoclonal antibodies including anti-CD99 (BD Biosciences, **supplementary figure 1**) on BD FACSLyric™ flow cytometer (BD Biosciences). Therefore, 100 µl EDTA blood was stained for 15 min at room temperature in the dark with monoclonal antibodies (BD Biosciences). Subsequently, red blood cells were lysed with BD FACS^™^ Lysing Solution (BD Biosciences) for 10 min, washed with 2 ml PBS. The pelleted cells were re-suspended in 300 µl PBS for analysis.

### Statistical methods, definition of biomarkers

Peptides levels across groups as well as flow cytometry data were compared using the Wilcoxon rank sum test. False discovery rate was assessed by the method described by Benjamini and Hochberg [12].

## Results

### Proteomics

Urinary peptide data of patients with critical COVID-19 course (n=53) were compared to data of mild COVID-19 courses (n=66). The statistical analysis returned 593 urinary peptides that were significantly associated with COVID-19 severity. Among peptides de-regulated in critical COVID-19 fragments of different types of collagens (53%), alpha-1-antitrypsin, beta-2-microglobulin, CD99, fibrinogen, polymeric immunoglobulin receptor (PIGR), and protein S100-A9 were mostly found.

To assess similarity to kidney disease (KD) [5], the data was further compared to peptide changes observed in DKD, exemplifying CKD, acute kidney injury (AKI), and heart failure (HF) as control, not expected to display similarity to critical COVID-19 course. Proteomics data from the human urinary peptide database from 571 DKD patients were compared to data of 374 controls [13;14]. For each peptide, fold change in disease vs. controls was calculated. Similarly, the fold change was also assessed for HF, investigating 773 datasets from HF patients compared to 773 controls [15]. To define the changes observed in AKI, data from 94 patients with AKI (KDIGO grade 2 and 3) were compared to controls (KDIGO grade 0, n=348) [16]. To compare changes observed in DKD, AKI and HF with those in critical COVID-19 disease, regression analysis was performed for all 593 peptides associated with COVID-19 severity. The results are shown in **figure 1**. A highly significant positive correlation between change in peptide abundance in COVID-19 and DKD (**figure 1A**) with R^2^ of 0.41 was detected. A slightly lower yet highly significant association was observed in comparison with AKI (R^2^=0.30, **figure 1B**). In contrast, no credible relationship between HF and critical COVID-19 was detected (**figure 1C**). This substantial degree of similarity between critical COVID-19 and DKD is also evident from the heatmap shown in **figure 1D** (for non collagen protein fragments) and **figure 1E** (for collagen protein fragments), depicting the urinary excretion pattern of the 100 most significant (p-values < 3.0×10^−7^) critical COVID-19 associated peptides. Further to the similarities to KD, the reduction of CD99 and PIGR peptides is apparently exclusive in COVID-19 and not detectable in DKD, AKI and HF.

**Figure 1:**
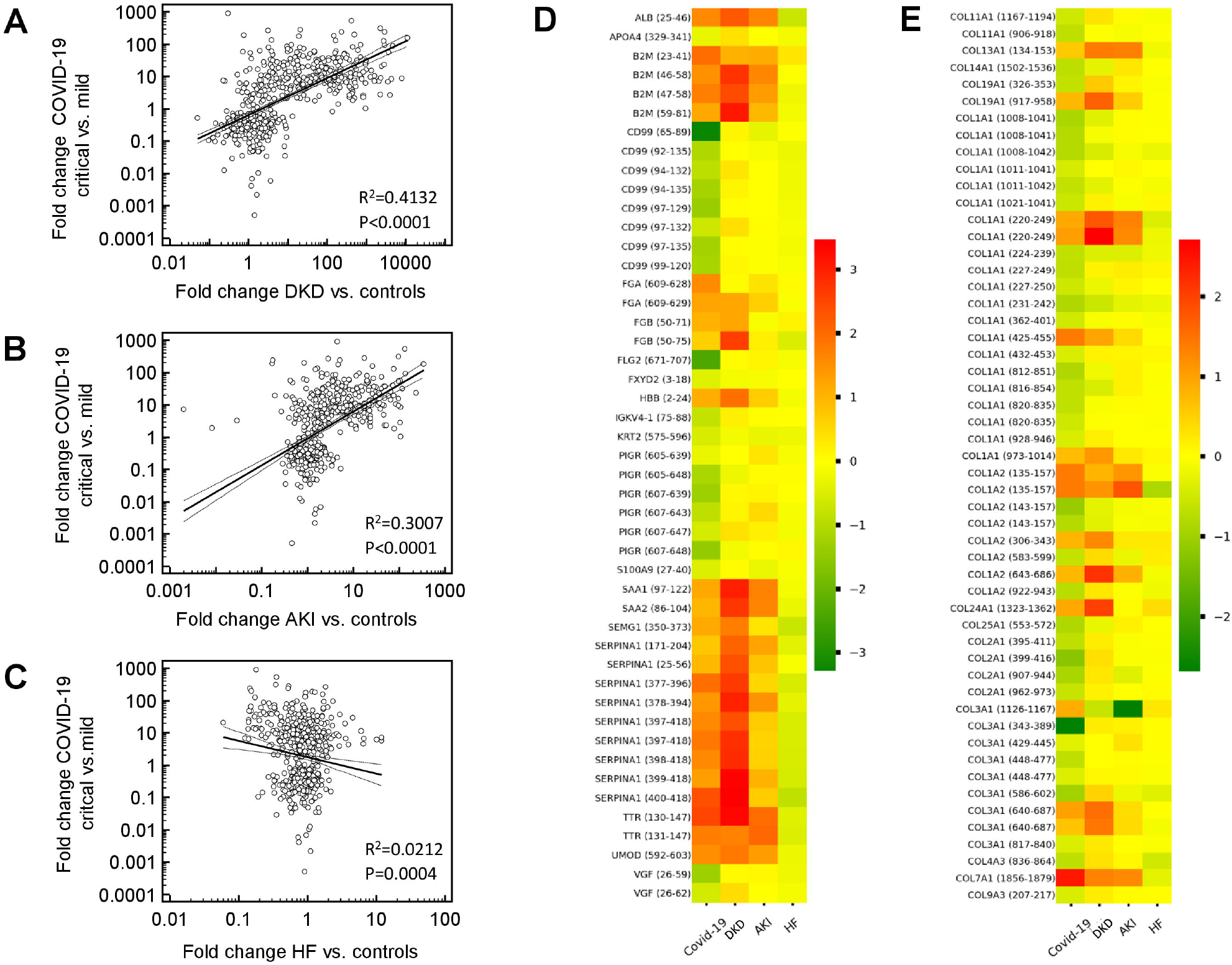
Association of changes observed in urinary peptides of critical COVID-19 patients and patients with diabetic kidney disease (DKD) acute kidney injury (AKI) and heart failure (HF). Regression plots of fold changes of all 593 critical COVID-19 specific urinary peptides (calculated critical vs. mild COVID-19) in comparison to changes observed in DKD patients (A), AKI patients (B) and HF patients (C) (calculated as disease vs. controls). Urinary excretion pattern of the 100 most significant (with p-values < 3.0×10^−7^) critical COVID-19 associated peptides is depicted as a heatmap for non collagen (D) and for collagen protein fragments (E).

### Flow cytometry

As a result of the observed reduction of CD99 peptides in critical COVID-19, we investigated CD99 expression on lymphocytes from COVID-19 patients, and from HIV-infected patients serving as controls. Since critical COVID-19 is associated with lymphopenia, the samples were separated into lymphopenic and non-lymphopenic. A significant decrease of CD99 expressing lymphocytes was detectable in patients with critical COVID-19, which was not detected in the control HIV-infected patients (**figure 2**).

**Figure 2:**
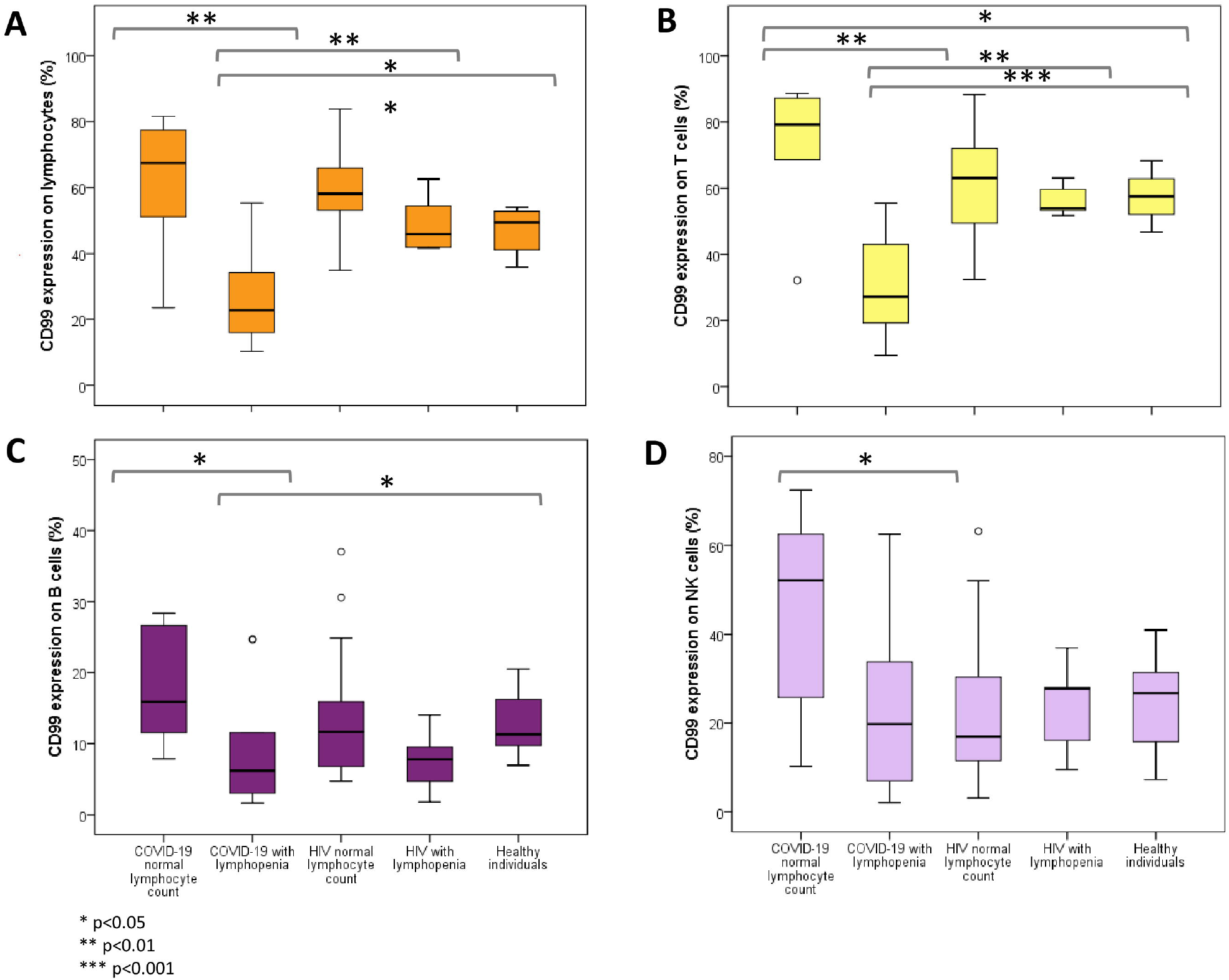
Relative CD99 expression of lymphocytes and of selected lymphocyte subpopulations. To evaluate CD99 expression on lymphocytes and lymphocytes subsets, lymphocytes (orange), T (yellow), B (dark purple) and NK cells (light purple) were selected by positivity of CD45, CD3, CD19, and CD16/56, respectively, using gating strategy depicted in supplementary figure 1. *p<0.05, **p<0.01, ***p<0.001

## Discussion

Changes in urinary peptides are specific for distinct pathologies and allow discrimination of different disease aetiologies as demonstrated for CKD [13], but also enable definition of shared changes displaying frequently involved processes like fibrosis or inflammation. The most prominently affected peptides in critical COVID-19 are collagen fragments, confirming previous observations describing the deregulation of collagen homeostasis [17]. Reduction of collagen peptides observed in KD as well [14], indicating molecular similarity between these diseases. Interestingly, the most prominent collagen fragments showed stronger decrease in critical COVID-19 patients compared to KD patients (**figure 1**). Similarities at molecular level between KD and critical COVID-19 patients were also observed in the regulation of alpha-1-antitripsin, in both cases up-regulated in urine, likely the result of increased degradation. Of note: Alpha-1-antitrypsin deficiency is described as major risk factor for critical COVID-19 [18].

Certain changes in urinary peptides appear unique for COVID-19. The exclusive reduction of CD99 peptides may indicate reduction of endothelial integrity and interference with transendothelial migration of monocytes, neutrophils, and T-cell recruitment. Loss of CD99 is expected to compromise tight junctions, possibly resulting in exposure of collagen, which would lead to (micro)thrombi. Analysis of lymphocyte subsets confirmed the reduction of CD99 in the group of critical COVID-19. Similar to proteomics data, in mild COVID-19 we observed an increase of CD99 expression on lyphocytes in comparison to healthy individuals, while in critical COVID-19 a large decrease of CD99 was observed.

Like CD99, PIGR has not been investigated in the context of COVID-19. This is the first report of an association of this protein with COVID-19 severity. PIGR was reported reduced in chronic obstructive pulmonary disease [19], a risk factor for COVID-19 severity. Further investigations of PIGR in COVID-19 patients via e.g. immunohistochemistry appear justified to confirm our observations.

The data suggest that a critical course of COVID-19 is associated with molecular changes typically present in KD, but not in HF. These observed changes may reflect endothelial damage with similarity to chronic kidney/microvascular disease. The data are supported by the fact that KD is an established risk factor for severe COVID-19 [4]. In contrast, based on our and others data, relevant association of HF with COVID-19 severity cannot be observed [20]. The data further indicate that therapeutic approaches based on improving endothelial integrity and preventing progressive endothelial damage may be beneficial in the management of critical COVID-19.

## Data Availability

All data contained in this manuscript will be made available upon request.

## Acknowledgments

The study was supported by the German Federal Ministry of Health acting upon a decree from the German Federal Parliament and by the DEFEAT Pandemics platform of the Federal Ministry of Education and Research (BMBF) to TW.

## Conflict of interest

HM the co-founder and co-owner of Mosaiques Diagnostics. JS, TH and AL are employees of Mosaiques-Diagnostics GmbH.

## Figure legends

**Supplementary Figure 1: Representative cytometric analysis by an 8-color panel including CD99**. A) Healthy patient with a lymphocyte count of 2800/ µl. B) Covid19 patient with lymphocyte count of 500 per µl. CD99 expression was measured on total lymphocytes, T, B and NK cells. As gating reference for CD99 relative expression, neutrophils served as negative-staining and monocytes as positives-staining populations. Mono = monocytes, Neutr = neutrophils, Lymph = lymphocytes.

**Supplementary Figure 2: Overview of immune perturbation in Covid-19**. Multiparametric flow cytometry analyses on fresh whole blood after red blood cell lysis characterizing lymphocytes and lymphocyte subsets in Covid19 patients with (n=6) and without lymphopenia (n=12), HIV infected individuals with normal (n=17) and lymphopenia (n=5). Lymphocytes (A), T cells (B), cells (C) and NK cells (D) were considered. As expected, we observed a significant reduction of lymphocyte subsets in individuals with lymphopenia in comparison to controls (p<0.005).

## Notes

### Author Declarations

This study was approved by the local ethics committee (Saxon Chamber of Physicians, registry number #EK-BR-88/20-1)

