## Supplementary figures and images for "Critical COVID-19 represents an endothelial disease with high similarity to kidney disease on the molecular level"

### supplemental figures 1 and 2

#### Slide 1
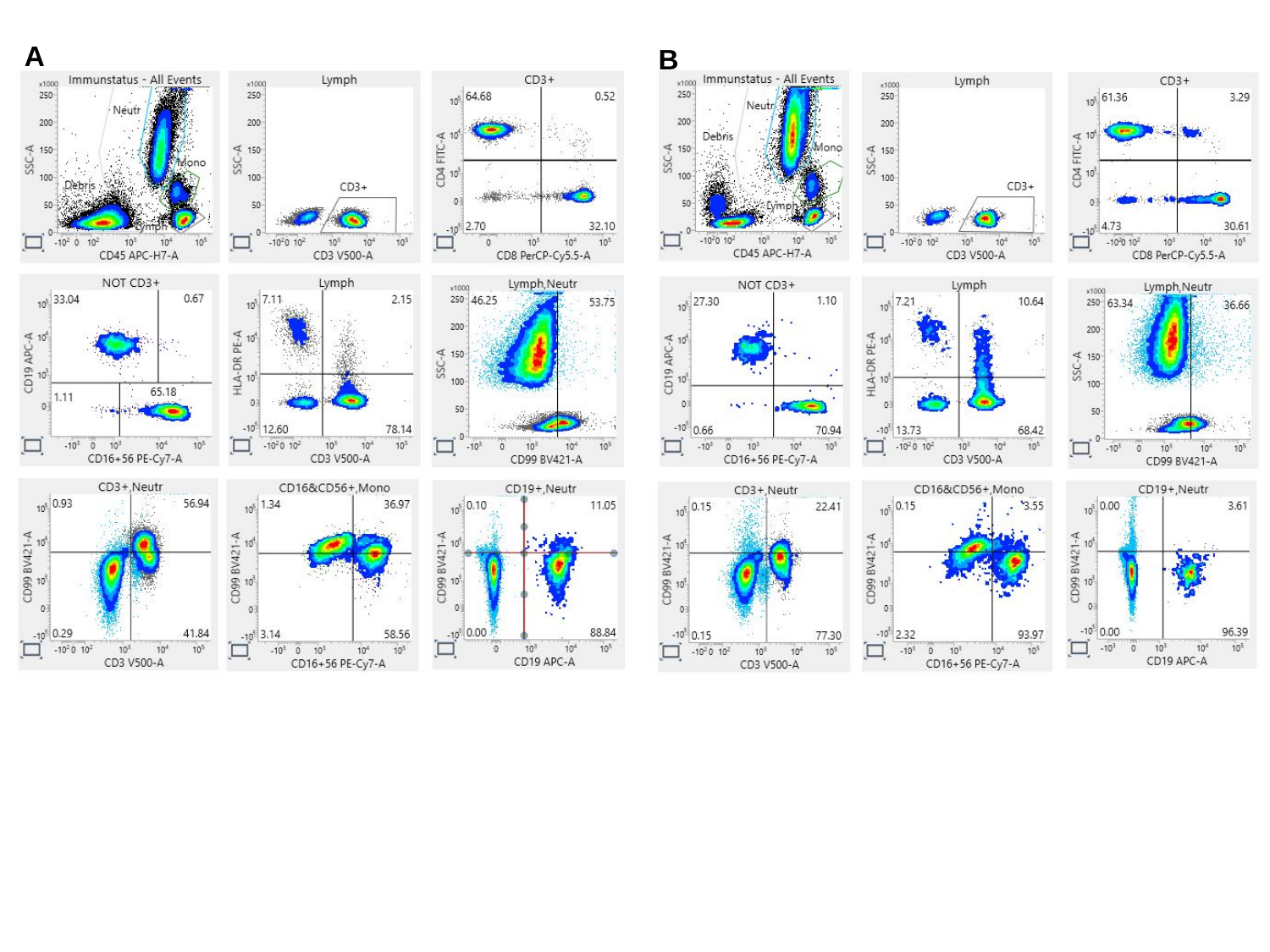

A
B

#### Slide 2
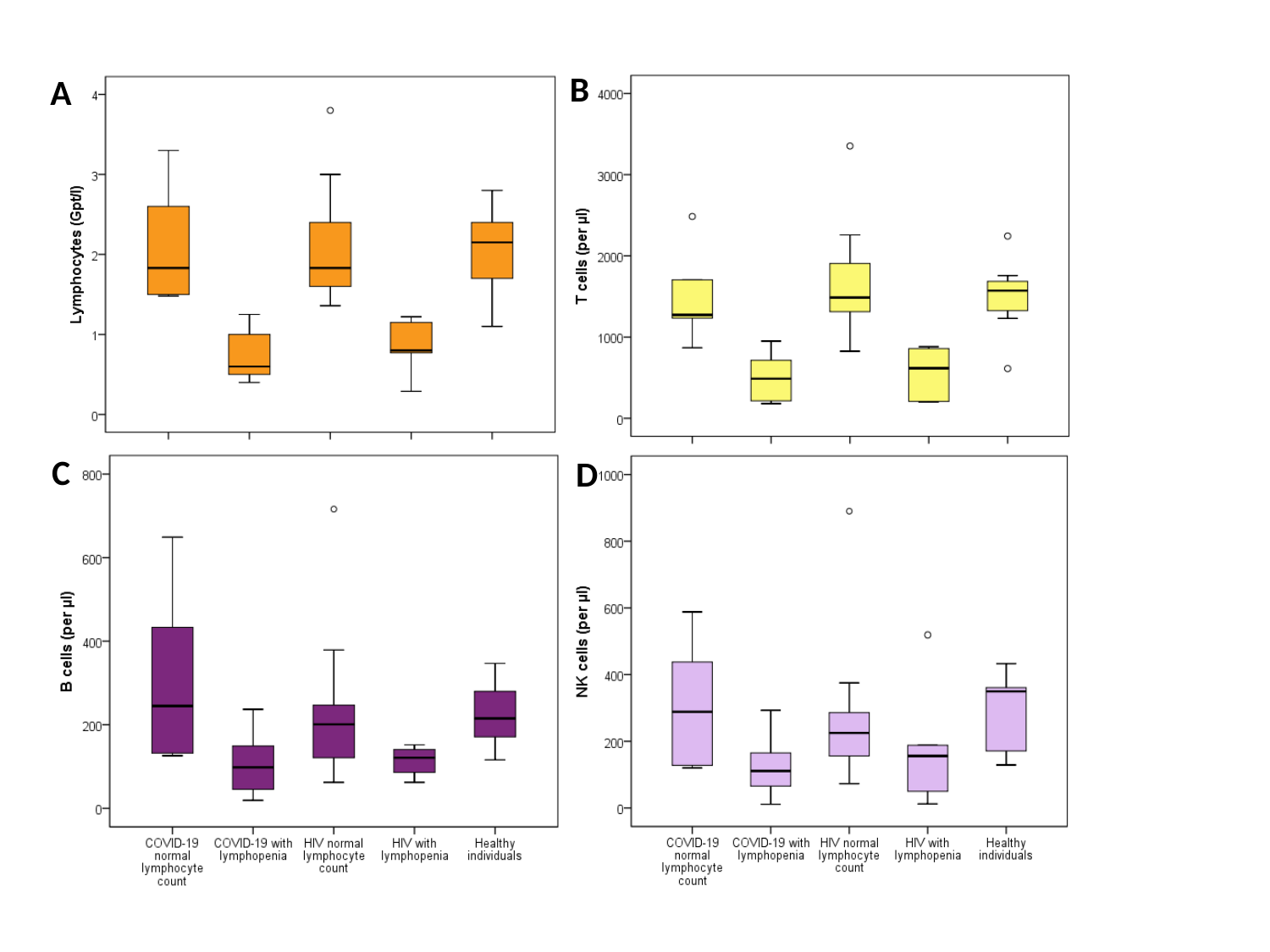

B
A
C
D
